# Frequency, Dynamics, and Duration of Fecal Shedding in SARS-CoV-2 Infected Individuals, a Scoping Review

**DOI:** 10.1101/2025.04.29.25326628

**Authors:** S Abunijela, T Greiner, W Haas, R Kerber, P Pütz, A Schattschneider, J Schumacher, U Buchholz

**Affiliations:** Department of Infectious Disease Epidemiology, Robert Koch Institute, Berlin, Germany

## Abstract

To estimate illness incidence or prevalence from wastewater data, modeling approaches may benefit from incorporating fecal shedding parameters. We systematically searched PubMed and a public repository on shedding data and included 33 studies meeting at least one of our objectives. Among 32 studies, the proportion of SARS-CoV-2-infected individuals with detectable virus in stool ranged from 18–100%, with a pooled estimate of 54% (95% CI: 52–56%). Stratification by four clinical severity categories, ranging from asymptomatic to critically ill, showed no significant differences among categories (p-value = 0.49). The proportion of individuals with detectable SARS-CoV-2 RNA in stool was higher in children (61%) than in adults (53%; p-value = 0.02). In half of the individuals who initially shed the virus in stool, it remained detectable for an estimated 22 days post-symptom onset. Three studies documented viral load kinetics, indicating a peak between days 3 and 9. Twenty-five studies reported maximum shedding durations ranging from 2 to 12 weeks. Our review presents a summary of the frequency, dynamics, and duration of SARS-CoV-2 shedding in stool and may serve as a valuable foundation for modeling efforts involving fecal shedding indicators.

## INTRODUCTION

SARS-CoV-2 transmission occurs primarily through respiratory pathways [1], while other transmission routes are of minor importance [2]. Nevertheless, many SARS-CoV-2-infected individuals release virus or viral fragments in their stool, enabling reliable detection in wastewater [3,4]. This has led to the widespread establishment of monitoring SARS-CoV-2 through wastewater surveillance systems.

Several studies have used fecal shedding data in combination with other information to predict SARS-CoV-2 incidence or prevalence from wastewater data [5,6]. These models rely on fecal shedding parameters to link the number of infected individuals with the amount of viral RNA detectable in wastewater. However, many studies have made assumptions or used shedding parameters from individual papers, such as the proportion of infected individuals who shed the virus, the timing of peak shedding, and the duration of shedding data derived from other clinical data [7]. Therefore, it is crucial to make shedding parameters available that integrate the current knowledge to refine wastewater-based models.

The average shedding of SARS-CoV-2 RNA in stool can be formally quantified using the following parameters:

1. The proportion of patients with diagnosed SARS-CoV-2 infection who have detectable viral RNA in stool.
2. Decline over time – how fast does the positivity rate (PR; proportion positive) of patients with initially detectable viral RNA in stool decline over time and after symptom onset.
3. Viral load kinetics – the timing of the magnitude of viral RNA in stool; and: what is the temporal sequence of the shedding peak in stool and respiratory samples, respectively?
4. Shedding duration – the maximum duration of fecal viral shedding time (VST).

Several reviews have addressed some of these questions and parameters [3,8–12]. However, some reviews did not apply standardized criteria for specific factors (e.g., requirement of systematic approach to stool sampling, considering the duration of follow-up, etc.). Additionally, some did not address key limitations of the underlying studies. Most reviews have made statements about the proportion of patients (or persons with SARS-CoV-2 infections) of whom viral RNA can be detected in the stool [3,9–12], and the maximum duration of virus in stool [3,11,13–15]. Two studies tried to characterize the dynamic aspects [6,16], and none has covered all aspects named above. In addition, the use of specific population subsets or differing methodologies of the underlying studies has led to inconsistent findings, particularly regarding how long individuals shed viral RNA in stool.

Our review aims to synthesize available evidence on the following parameters:

1. proportion of persons infected with SARS-CoV-2 who shed viral RNA in the stool
2. (a) among persons with fecal shedding: decline of positivity rate over time, and (b) dynamic course of viral RNA in stool over time
3. maximum duration of fecal viral shedding.

## Methods

### Information Sources and Search Strategy

We searched PubMed using a combination of MeSH terms and keywords linking the three categories: population (SARS-CoV-2-infected individuals), sample material (stool), and parameters (including positivity rate, kinetics, and viral load) using the AND operator, (Appendix; Table A1). Additionally, we screened a public repository on shedding data [18] to identify further relevant studies.

### Inclusion and Exclusion Criteria

We included studies in which SARS-CoV-2 infections were confirmed via positive respiratory swabs prior to stool sample collection. Also, we included only studies that started to take stool samples of participants during the first week after symptom onset. Only studies that collected at least three stool samples in majority of participants were selected.

To calculate the proportion of SARS-CoV-2-infected individuals in whom SARS-CoV-2 was also detected in stool (objective 1), only studies that reported this indicator based on the number of participants, rather than the number of samples, were included. To estimate the proportion of SARS-CoV-2-infected individuals with positive stool (trend of the PR) over time (objective 2a), studies that involved SARS-CoV-2-infected individuals, who regularly provided stool samples over a period of time, were included. To calculate the shedding dynamic (of the PR (objective 2a) or the viral load (objective 2b)), only studies that specified the day of sample collection with respect to symptom onset were included (and not, for example, with respect to hospital admission), as well as studies that incorporated SARS-CoV-2-negative stool samples in their proportion of patients with fecal shedding or in their analysis of viral shedding kinetics. To calculate the maximum VST of the detectable virus in stool samples (objective 3), only studies that provided duration of VST, with a minimum follow-up period (at least 2 weeks), in time after symptom onset were included.

We excluded studies with unclear methodologies, such as studies that were lacking information on the number of stool samples taken per patient, as well as studies with vague patient selection criteria. Additionally, during the screening process, we disregarded studies that focused on animals and those that were not written in English or German.

### Selection of Studies

We imported the identified studies to EndNote for de-duplication and transferred the remaining studies to Microsoft Excel (Microsoft Corporation, Redmond, WA, USA). One author (S.A.) reviewed the titles and abstracts of the identified studies according to the inclusion and exclusion criteria and classified them as preliminarily included, excluded, or uncertain. Studies that were initially classified as ‘uncertain’ were discussed with a second author (U.B.) until all conflicts were resolved. Full texts were then carefully examined by (S.A.) and again reclassified as included, excluded, or uncertain, and remaining uncertainties were resolved through discussion with (U.B.).

### Data Extraction

For each included study, we systematically extracted the following details: author(s), journal, publication year, title, main findings, including the proportion of infected individuals with shedding, PR over time, shedding kinetics, and duration of shedding. One reviewer (S.A.) extracted the data while a second reviewer (U.B.) checked the accuracy to ensure consistency and reliability.

### Calculation of key parameters

For the three study objectives, we calculated the proportion of infected individuals that also shed virus in the stool, the PR over time, the kinetics of the viral load, and the VST. Details and the related statistical analysis are summarized in (Table 1).

**Table 1.**
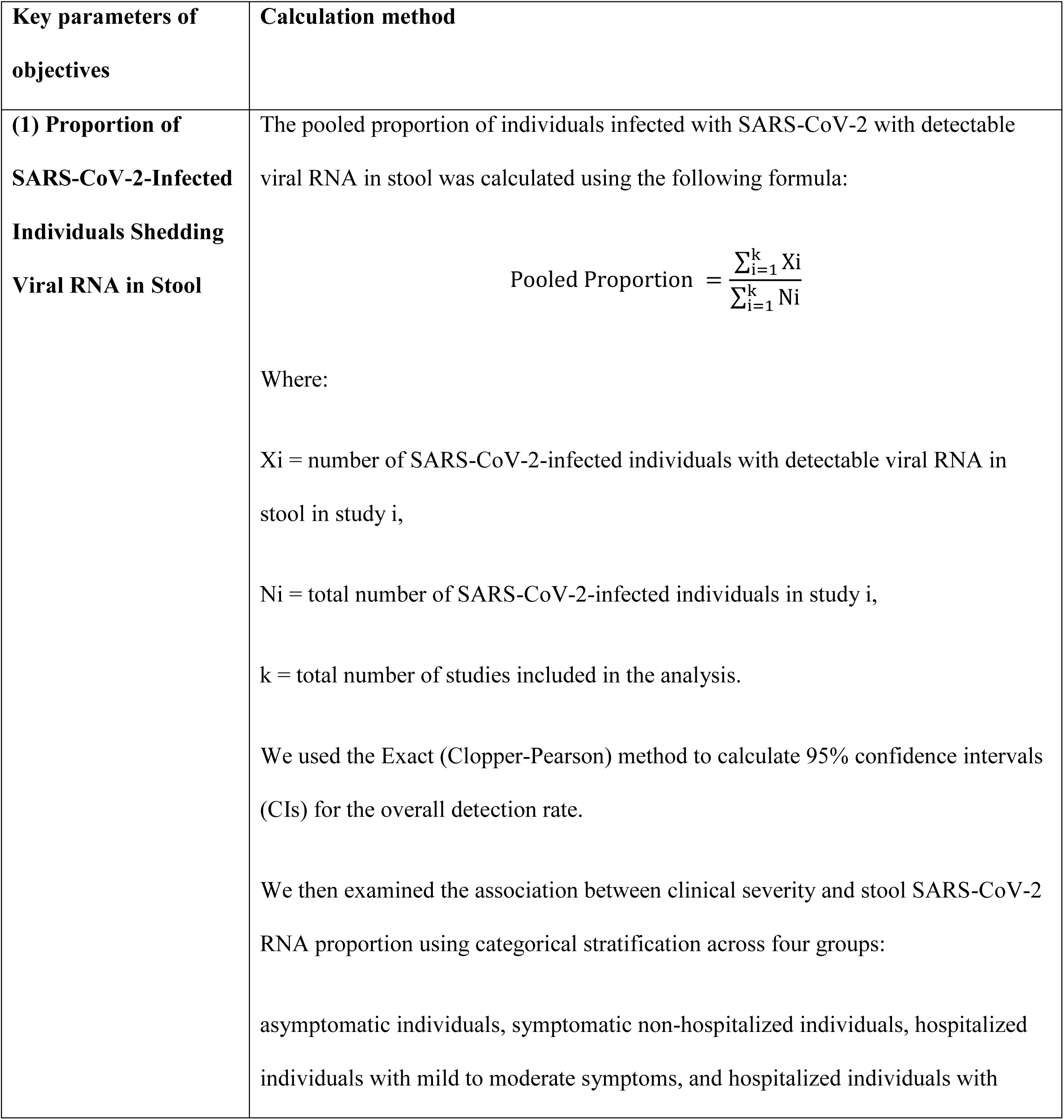
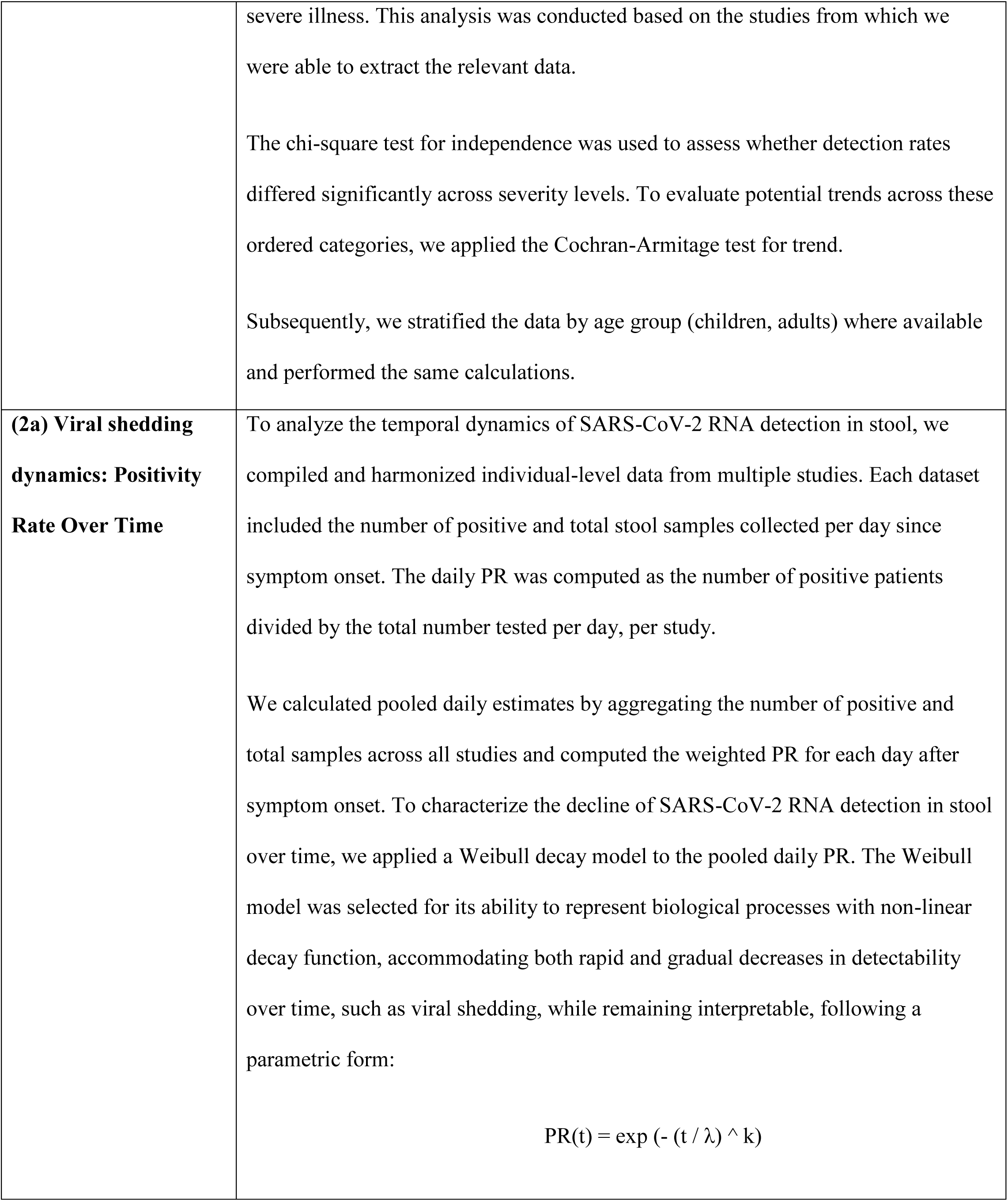
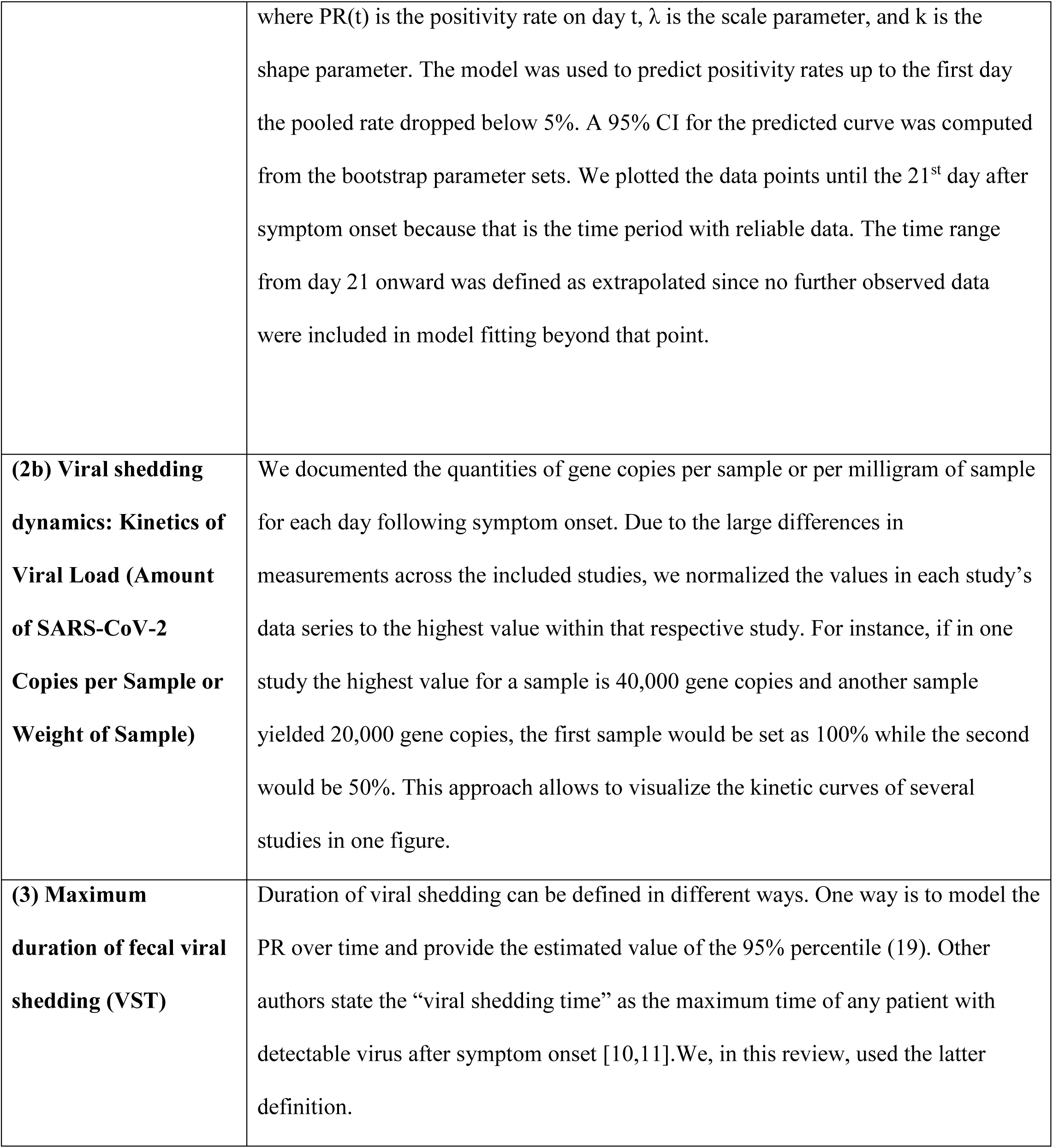
Summary of the calculation of key parameters.

## Results

The PRISMA flow chart [20] outlines how included studies are chosen, as illustrated in (Figure 1). Out of the 644 identified studies, 33 primary studies were included in this review. Most studies were conducted between January 2020 and August 2021, with two later studies in February 2021– January 2022 [19] and 2023 [21]. Participants of studies exclusively conducted in 2020 were not vaccinated, or vaccination was not reported. Similarly, participants of studies totally or partially conducted in 2021, i.e. when vaccines had become increasingly available, were primarily not vaccinated or vaccination was not reported, with few exceptions (Appendix; Table A2). Notably, the study that was conducted in 2023, involved fully vaccinated participants, mostly without booster, with a 100% detection rate of SARS-CoV-2 RNA in stool [21].

**Figure 1.**
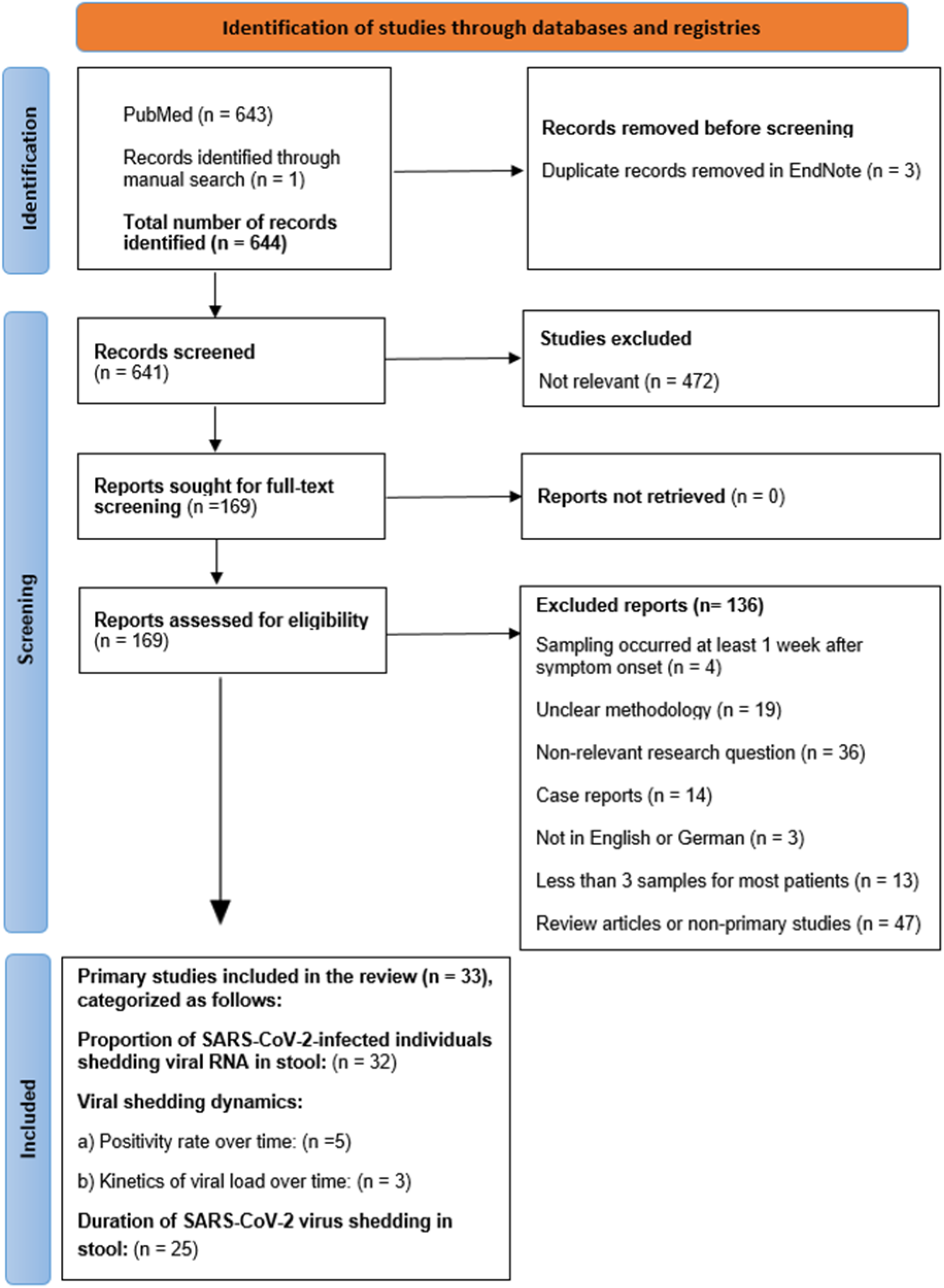
PRISMA flow chart depicting the screening process for filtering suitable primary studies for the review.

### Objective 1: proportion of persons infected with SARS-CoV2 who shed viral RNA in the stool

Out of the 33 analyzed studies, 32 provided data on the proportion of SARS-CoV-2-infected individuals with detectable viral RNA in their stool (Figure 2). The studies included a range of participants varying from 5 to 280, (Median = 45). Furthermore, the proportion of SARS-CoV-2 infected persons with fecal shedding varied from 18% to 100%, with a pooled proportion of 54% (995 of 1847 patients; 95% CI = 52%–56%, Figure 3A).

**Figure 2.**
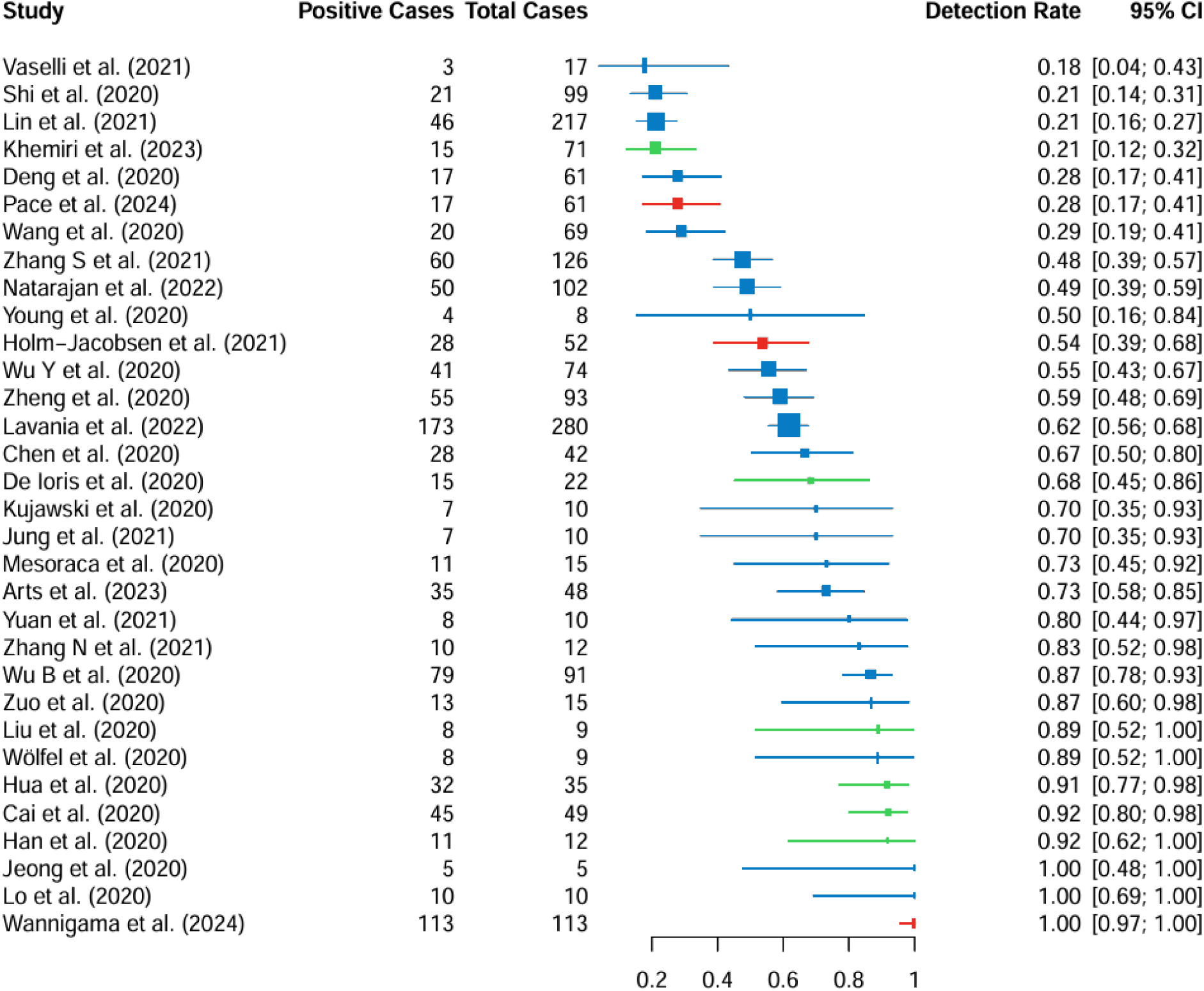
Proportion of individuals with SARS-CoV-2 infection (total cases) in whom viral RNA was detected in stool samples (positive cases): studies highlighted in green included only children, red included both children and adults, and blue included only adults. Squares represent the effect estimate for each study (the proportion of SARS-CoV-2 infected individuals with fecal shedding of viral RNA), with larger squares indicating larger sample sizes. Note: This figure provides a descriptive summary of detection rates across studies without meta-analytical models. The Exact (Clopper-Pearson) method was used to calculate 95% CIs.

**Figure 3.**
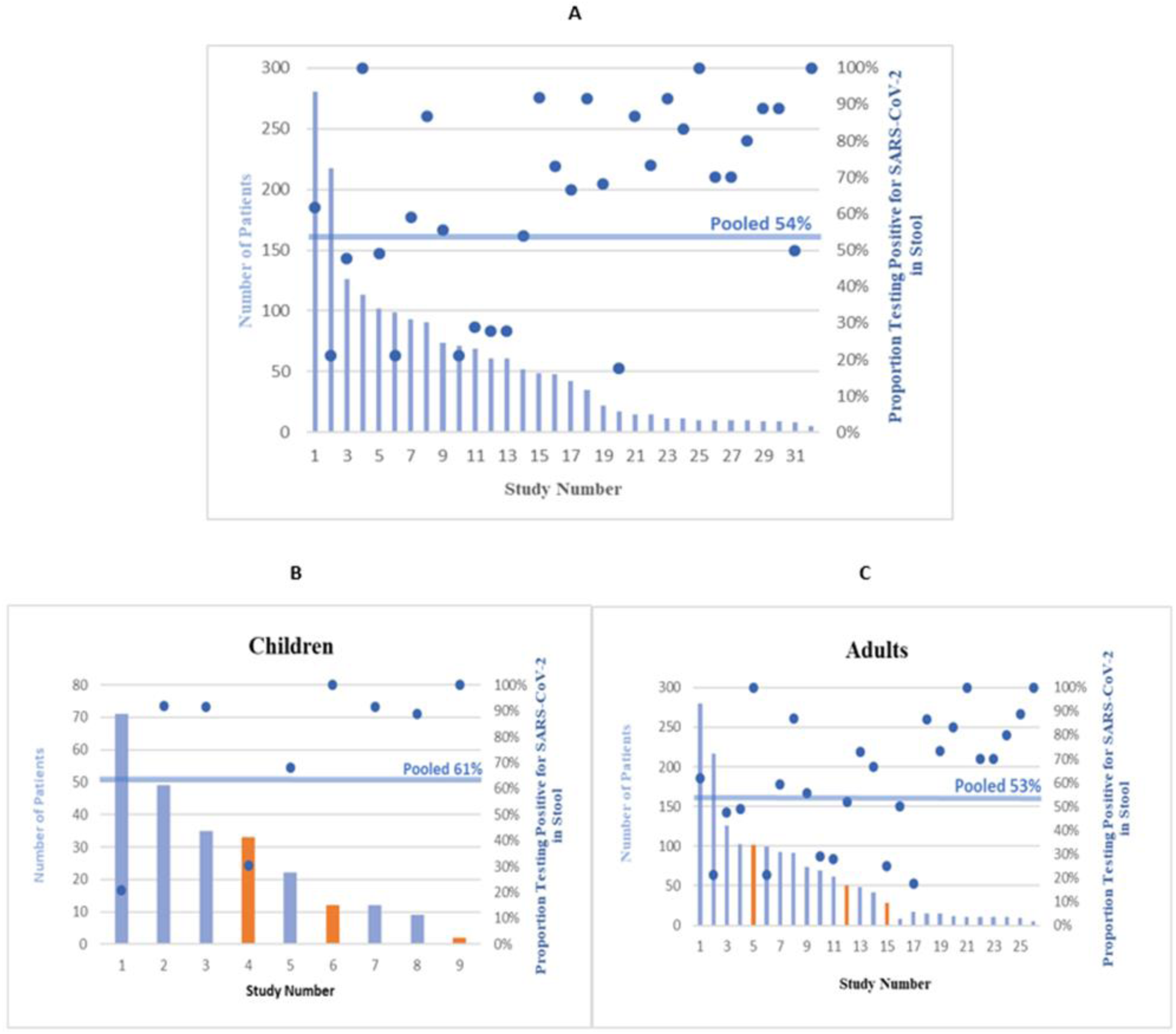
Proportion of patients with detectable SARS-CoV-2 in stool: (A) all studies, (B) studies only in children (Of these, 6 targeted only children [19,22–26] and 3 studies included children and adults [21,27,28]), and (C) studies in adults (23 of these addressed adults only [7,29–50]). The x -axis displays the number of the studies. The left y -axis (relating to the bars) represents the quantity of patients per study and the right y -axis (relating to the blue dots) indicates the proportion of SARS-CoV-2-infected individuals testing positive for the virus in stool. The pooled proportion is displayed by the horizontal blue line. The orange bars in chart (b) and (c) represent the studies that involved both children and adults that contributed data for the respective age categories.

Pooled proportions were higher in children (61%) compared to adults (53%) (p-value = 0.02), suggesting a significant difference in detection rates between the age groups. Detection rates stratified by clinical severity ranged from 35% in asymptomatic individuals to 42% in those with severe illness, without significant differences (p-value = 0.49; p-value for trend = 0.16). Although most studies are shared across both subgroupings, stratification by age and by clinical manifestation emphasizes different aspects of the population. Age-based stratification includes all reported individuals, regardless of symptom status or illness severity. Detailed subgroup results are shown in Table 2 and Figures 3A–3B.

**Table 2.**
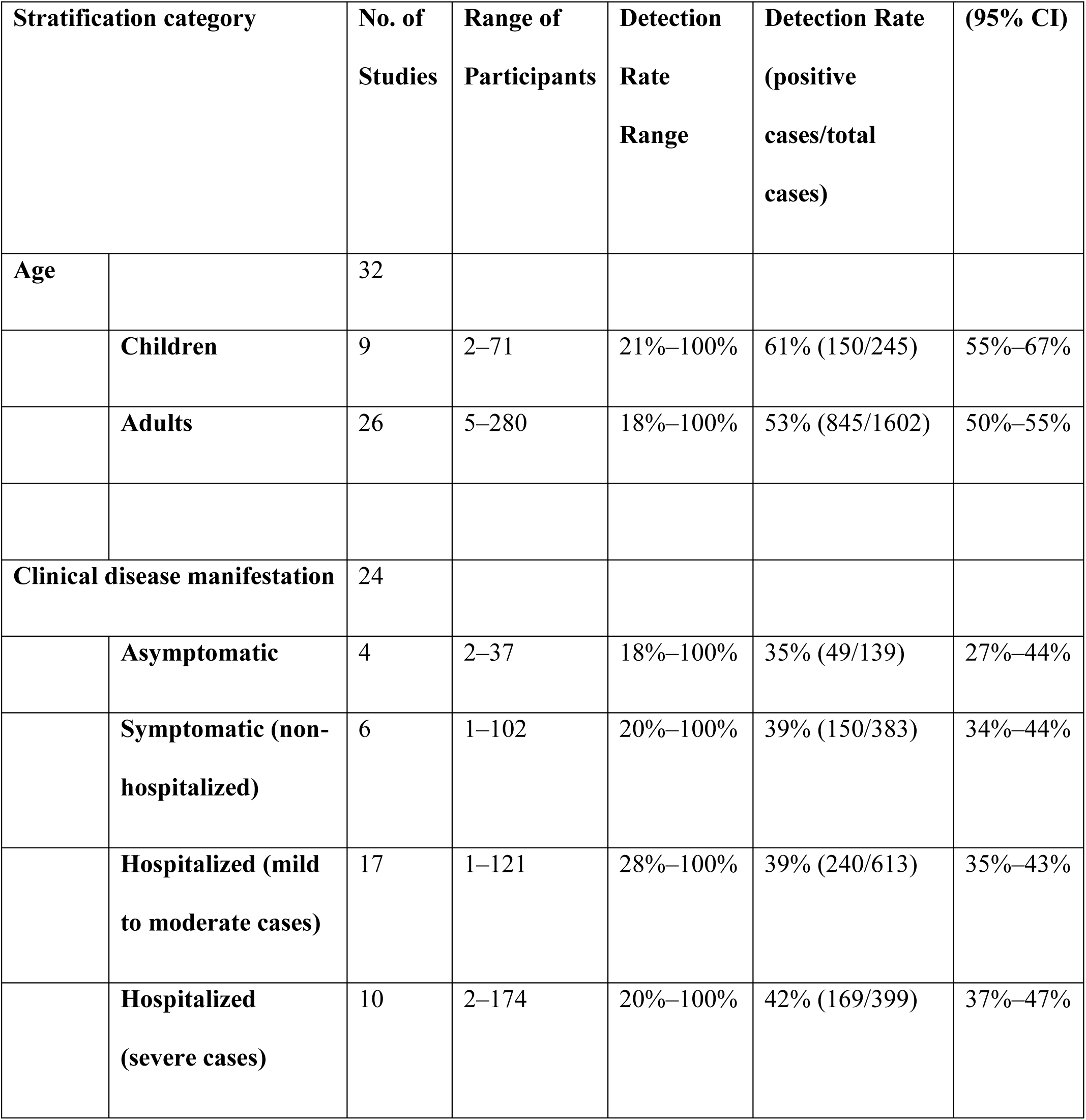
Proportion of SARS-CoV-2-infected Individuals Shedding Viral RNA in Stool by Subgroup.

### Objective 2a: Viral shedding dynamics: positivity rate over time

For this analysis, we included data from five studies (466 stool samples through day 21), (Figure 4). The pooled positivity rate exceeded 75 % in the first week. According to the trend analysis, 50% of the people, who initially excreted SARS-CoV-2, still excreted SARS-CoV-2-positive stool at day 22 after symptom onset, i.e. over a period of around 3 weeks. The Weibull model predicted a drop below 5 % at day 65. Study-level variability was visible, particularly in early and late time points, but the overall pooled trend was robust.

**Figure 4.**
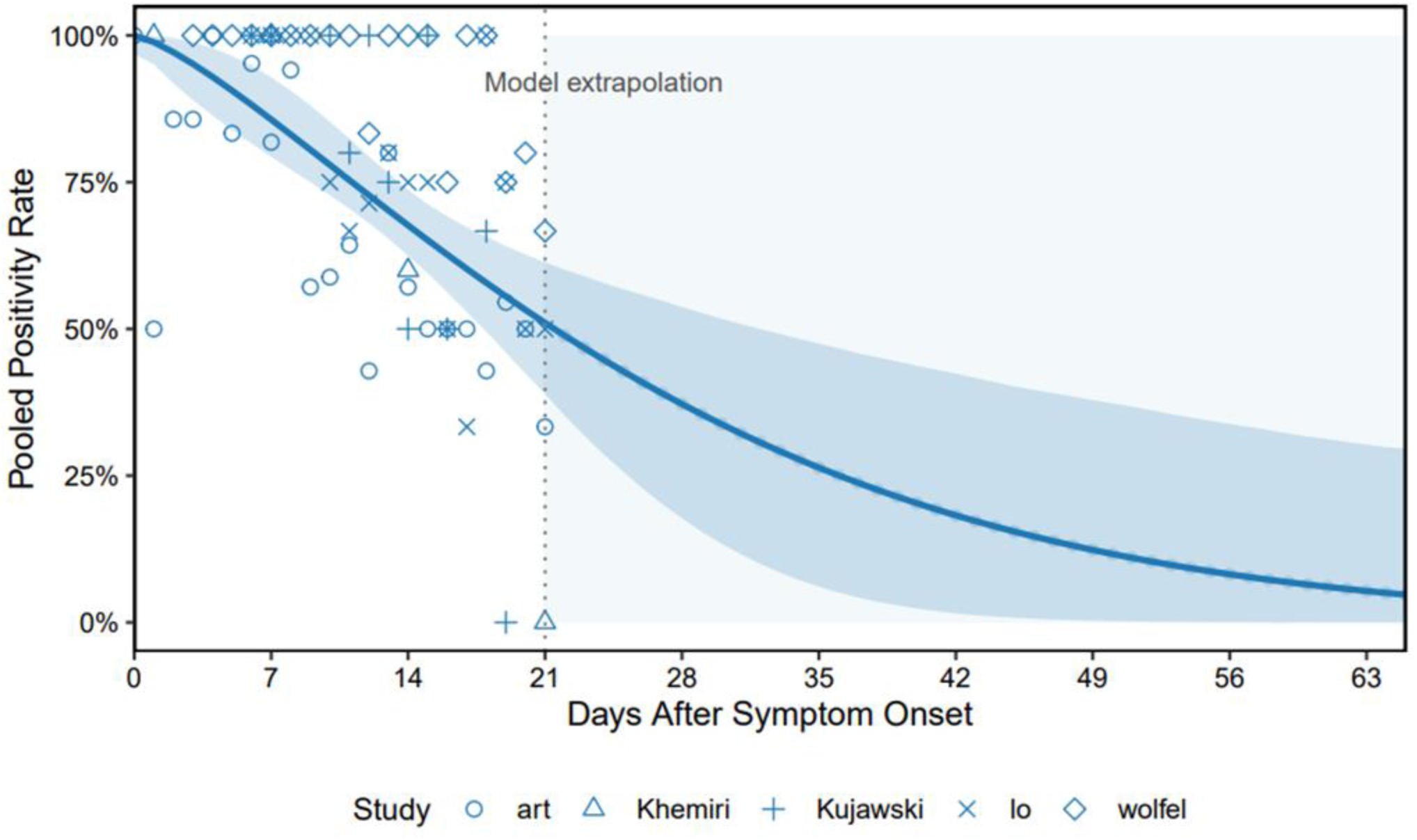
Pooled proportion of SARS-CoV-2 RNA-positive stool samples by day since symptom onset across multiple studies 5 studies [7,19,30,32,36]. X-axis shows days after symptom onset; Y-axis shows pooled PR. The observed PR from five studies are displayed as blue markers with distinct shapes. The solid blue line represents the fitted Weibull decay curve to the daily weighted PR computed from pooled data. The shaded blue ribbon depicts the 95 % CI derived from 5000 bootstrap replicates. A vertical dashed line at day 21 marks the end of empirical data; the pale-blue shaded area to the right denotes the extrapolation range.

### Objective 2b: Viral shedding dynamics: kinetics of viral load over time

We extracted the information regarding the amount of the viral load in stool from only three studies that provided robust data [7,21,36], (Table 3). Figure 5 shows normalized viral kinetics by days after symptom onset. For reference, viral load kinetics in the upper respiratory tract (URT) from a separate study are also displayed (green squares) [51], typically preceding the fecal kinetic curve by 3-10 days. Sharp peaks are noticed across the different SARS-CoV-2 variants included in each study, with peaks occurring at day 3 [36], 7 [7], and 9 [21], respectively.

**Figure 5.**
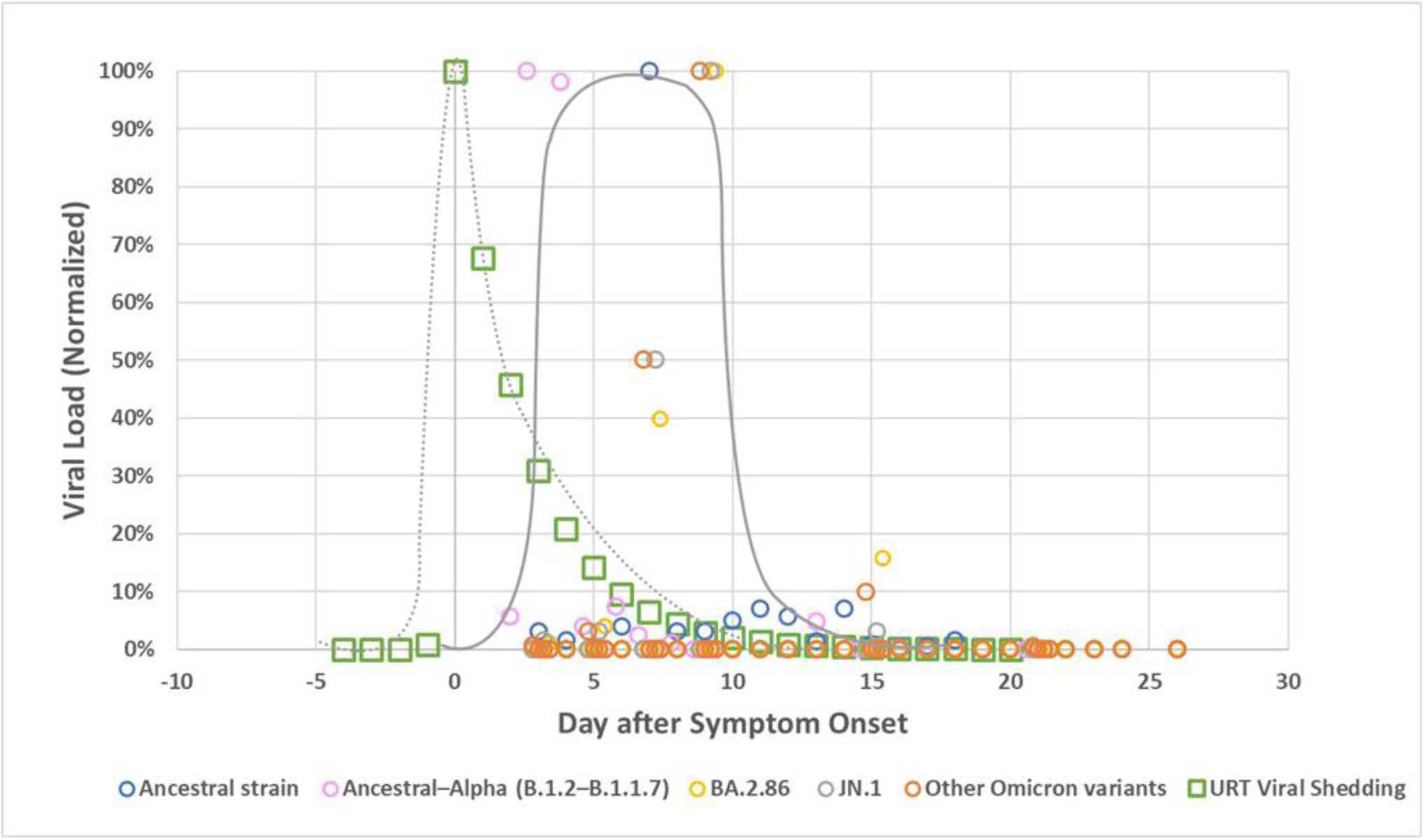
Normalized viral load in stool (expressed as a percentage of the maximum value within each data series in stool and upper respiratory tract (URT) samples from individuals infected with SARS-CoV-2, plotted by days after symptom onset. The circles represent stool viral load data derived from three studies [7, 21, 36]. Blue circles correspond to study [7] and reflect the ancestral strain. Pink circles represent data from study [36] and include both ancestral and Alpha variants. Yellow, gray, and orange circles originate from study [21] and represent different Omicron variants. Green squares show URT viral data for comparison [51]. Solid and dashed gray lines represent non-parametric approximations of fecal and URT viral kinetics, respectively. These lines were included for visual comparison only and were not derived from formal statistical fitting. The y-axis presents normalized viral load values on a linear scale.

**Table 3.**
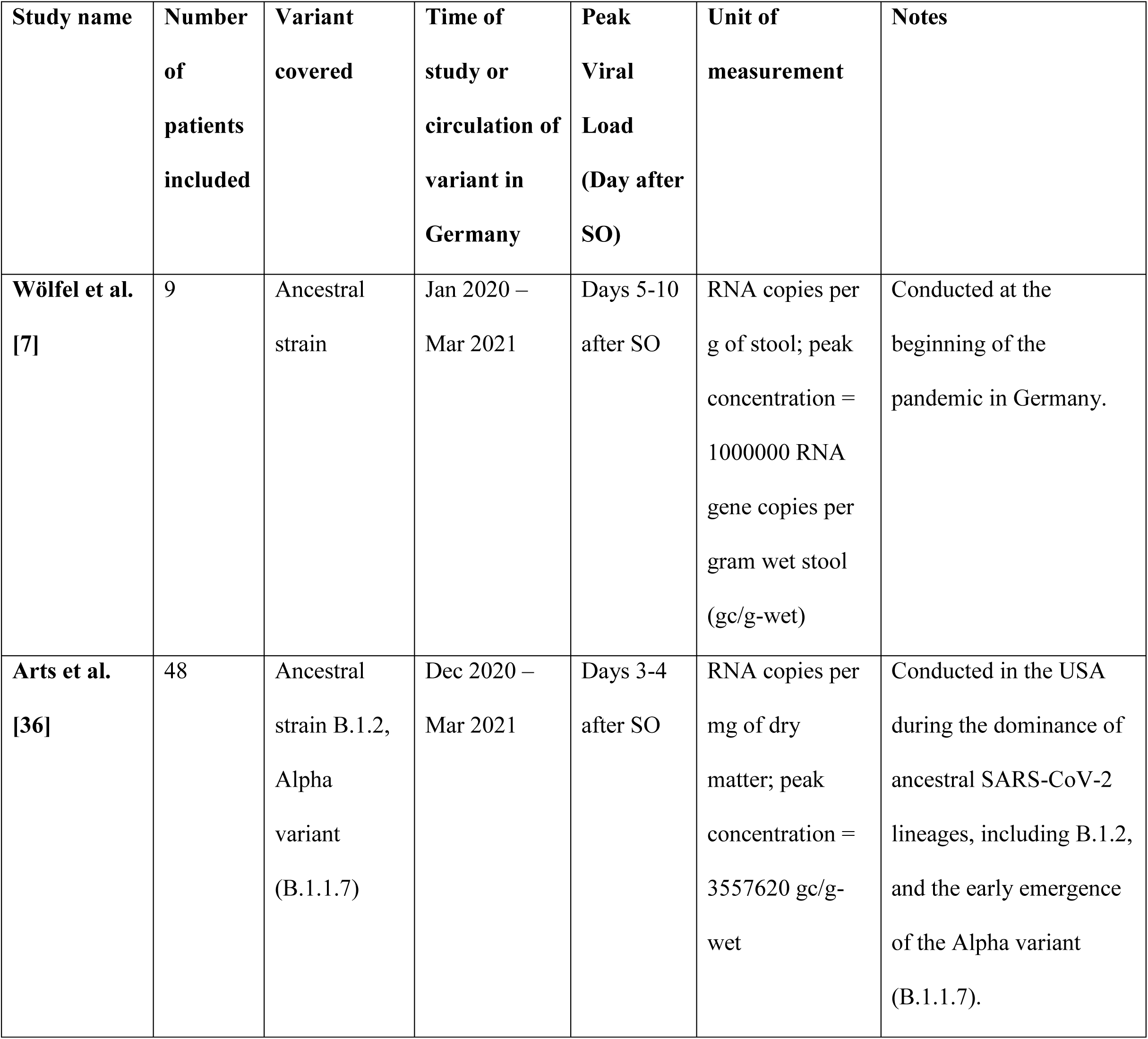
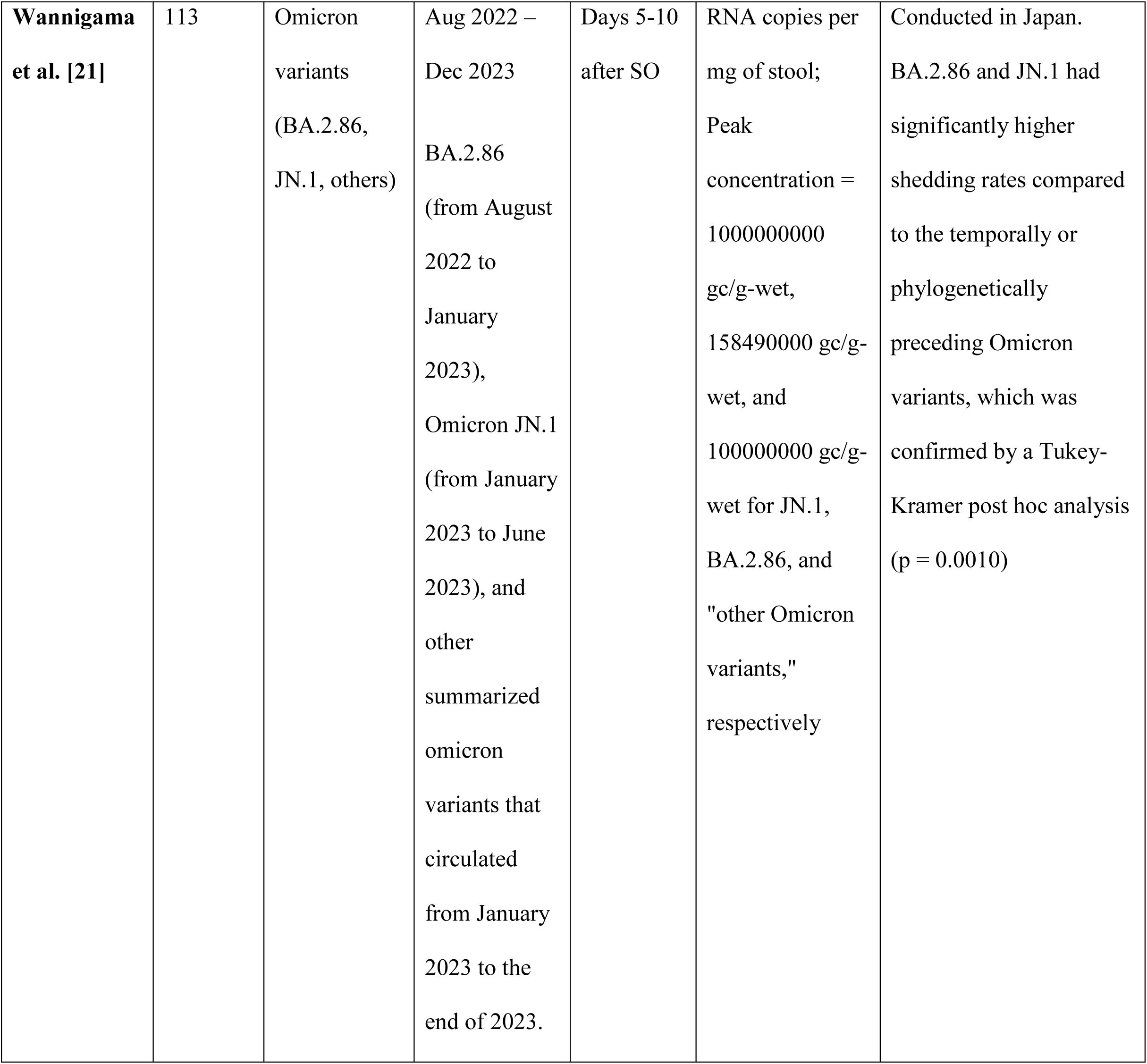
Comparison of Peak SARS-CoV-2 Viral Load Kinetics Over Time and Variants: Insights from Three Studies; SO= symptom onset. Note: Study [36] used dry weight, while the other two studies [7,21] used wet weight. To harmonize the units, we converted dry to wet weight based on a wet mass of 128 g/person/day, corresponding to a dry mass of 29 g/person/day [59].

Numerous studies have examined the temporal kinetics of virus shedding using Ct values [19,27,28,30–32,44,45,47–50], Kaplan-Meier plots [25,26,43], or Weibull -Regressions [17]. In studies that used Ct values and therefore did not report a direct quantitatively interpretable viral load, SARS-CoV-2 RNA was found to be detectable in almost all patients in the first week after symptom onset. After the second week of symptom onset, Ct values increased rapidly, indicating an exponential decrease in viral load. Hence, these studies support results of the three studies illustrated in (Figure 5).

### Duration of SARS-COV-2 virus shedding in stool

The duration of detection of SARS-CoV-2 RNA in stool among SARS-CoV-2-infected people that shed the virus at all has been measured in several studies. Some have stopped taking fecal samples in individual patients after obtaining one, or mostly two, negative samples. Several studies have followed up patients for the time period they were hospitalized, which favours obtaining samples from patients with more severe disease, whose stools were perhaps more likely to still be positive for a longer time than among patients with milder disease. Thus, most studies did not sample their entire patient cohort (or even close to entirety) systematically for the same extended period of time. Due to limitations in the available studies, we have refrained from calculating a summary value. The included 25 studies followed between 3 and 173 patients, (Median =13), (Table A3 (13 studies with systematical sampling) and Table A4 (12 studies without systematic sampling), (see Appendix)) (Figure 6). While the majority of studies reported shedding durations ranging from 2 weeks to 12 weeks, one exceptional study documented a considerably longer shedding period [42]. This study monitored 60 patients for 28 weeks (7 months), collecting an average of three stool samples per patient. After seven months, SARS-CoV-2 RNA was detected in 4% of samples, but it is not clear if these patients had provided stool samples and tested positive continuously throughout the entire seven months. Of the 25 studies, 20 provided a direct comparison of respiratory and fecal shedding. Among those, 17 studies found that shedding in the respiratory tract stopped before fecal shedding ceased [7,17,19,22,23,28,31–35,37,40–42,44,45], while three studies found that shedding in the respiratory tract lasted longer than in stool [26,30,49]. In (Figure 7), we recap the findings of this review schematically.

**Figure 6.**
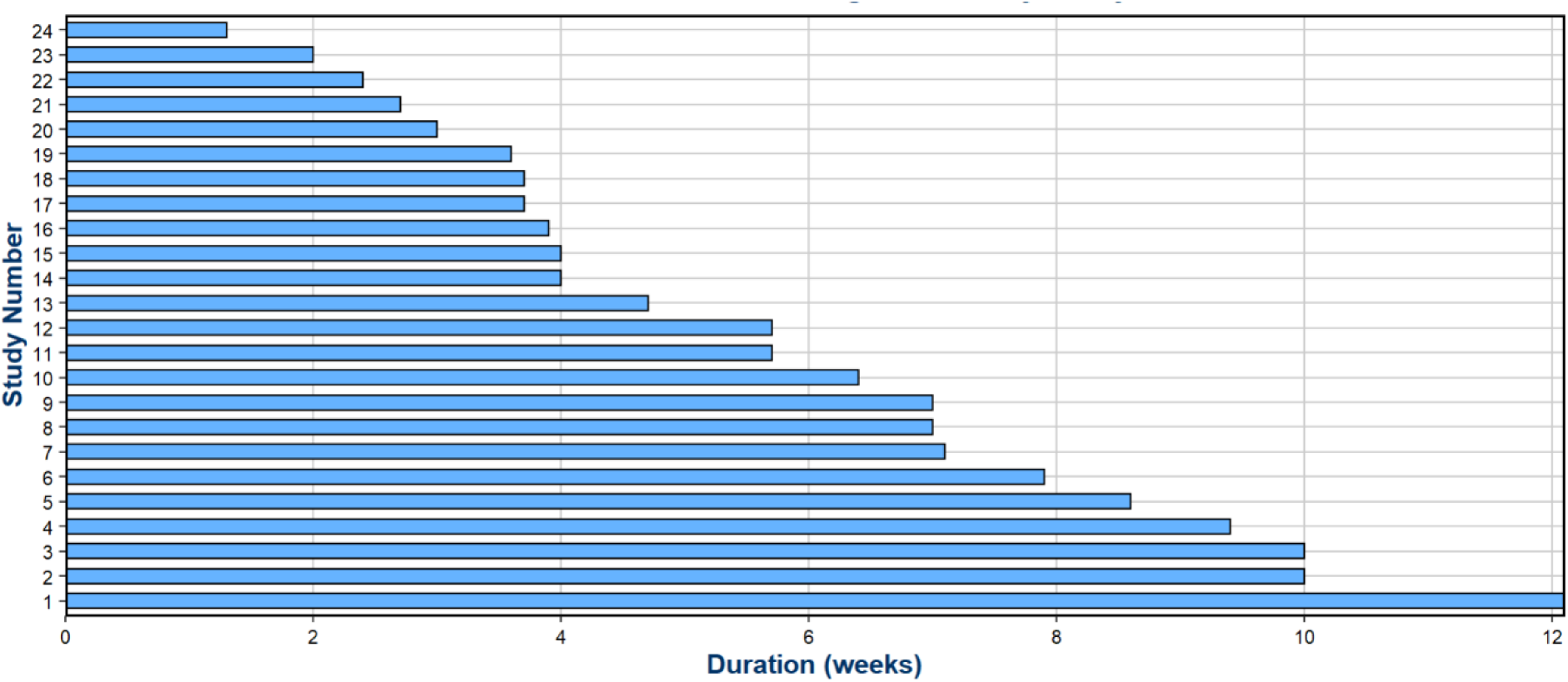
Maximum duration of SARS-CoV-2 stool shedding reported across the 24 studies. Note: one exceptional study [42] providing positivity rates up to 7 follow-up months is not considered here, for more details see the ‘Results’ section.

**Figure 7.**
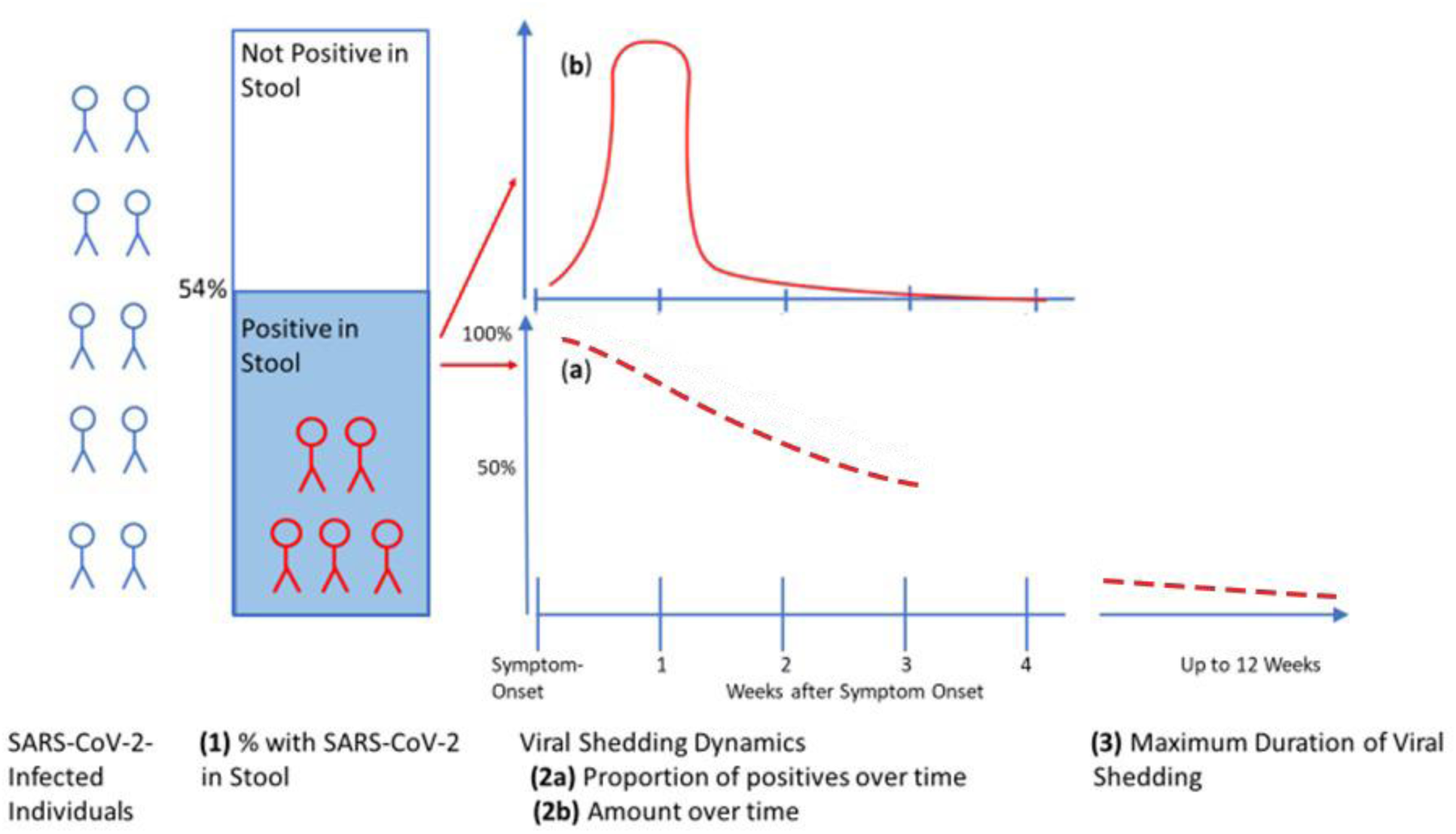
Idealized summary of the review findings on SARS-CoV-2 shedding in the stool of infected individuals. Note: Regarding the maximum duration of viral shedding, the result of one exceptional study [42], is not considered here, for more details see the ‘Results’ section.

## Discussion

This review aimed to examine the frequency, dynamics, and duration of SARS-CoV-2 RNA shedding in the stool of infected individuals. We found that approximately half of people infected with SARS-CoV-2 shed viral RNA in the stool. While significant differences were identified between age groups, no significant differences were found when stratifying by clinical severity. We estimate that among SARS-CoV-2-infected individuals who shed SARS-CoV-2 in their stool, about half of these do so for 22 days, with a peak viral load observed during the first or early second week after symptom onset. Maximum viral shedding time is reported to range between 2 and 12 weeks.

### Proportion of persons infected with SARS-CoV2 who shed viral RNA in the stool

Our estimated proportion of SARS-CoV-2 infected persons, who shed viral RNA in their stool (54%), is comparable to findings from four systematic reviews estimating between 48% and 55% [8,9,11,52], while three others reported proportions farther away (35% [10], 43% [14], and 66% [3], respectively). Differences between reviews may be attributed to variations in the inclusion criteria of the included studies. At any rate, the studies included in our review are remarkably heterogenous stemming from different study designs, patient populations, or methods used during the data collection process.

Further stratification by clinical severity revealed detection rates of 35% among asymptomatic individuals, 39% among symptomatic non-hospitalized individuals, 39% among hospitalized individuals with mild to moderate symptoms, and 42% among those hospitalized with severe illness. These differences were not statistically significant (chi-square test, p-value = 0.45), suggesting no clear link between symptom severity and stool-based viral RNA detection. A meta-analysis reported a higher proportion of fecal RNA detection in patients with more severe disease (60% vs. 47% for mild or moderate cases), although the between-group differences were also not statistically significant [53]. Similarly, another meta-analysis reported no significant difference in fecal RNA positivity between severe and non-severe cases (odds ratio = 2.009, p-value = 0.079) [54].

The majority of included studies, in our review, were based in clinical or hospital settings, which likely overrepresent individuals with more severe disease. Asymptomatic or mildly symptomatic individuals—who may not seek medical care or be routinely tested—are likely underrepresented.

While the lack of significant association in our findings does not rule out the possibility of a relationship between severity and viral shedding, factors such as the timing of sample collection could influence detection rates. For example, hospitalized patients may have their samples collected later in their illness, potentially leading to underestimation of shedding proportion.

Although we did not conduct a formal meta-analysis or funnel plot to assess publication bias, it is important to note that studies with low or negative detection rates may have been less likely to be submitted or accepted for publication, potentially inflating the overall positivity rate estimates. Future studies might recruit participants using community-based strategies, such as targeting households with a positive primary case, to obtain more accurate estimates with minimal bias and assess the true burden of stool-based viral RNA shedding across different clinical presentations.

The proportion of individuals with detectable SARS-CoV-2 RNA in stool was higher in children (61%) than in adults (53%; p = 0.02). Although two systematic reviews reported higher detection rates in children, they concluded that the difference was not substantial [10,14]. Three other systematic reviews and meta-analyses found considerably higher detection rates in children (86%– 89%) [55–57], compared to 54% in adults [4], although statistical significance was not discussed. This finding is noteworthy, as children have been reported to shed significantly less SARS-CoV-2 in respiratory samples compared to adults [51,58]. It remains unclear why viral traces cannot be detected in stool samples from many infected individuals. Possible reasons include the sensitivity of the detection methods, timing of collection of the stool sample, prevailing immunity at infection, or biological variation among infected hosts.

### Viral shedding dynamics: (a) positivity rate over time, after symptom onset

Our results indicate that half of the people who shed viral RNA in the stool will do so for about 22 days, which equals the median clearance reported in a previous systematic review [10]. Additionally, these findings align with another early systematic review by Morone et al. (2020) with a somewhat shorter median duration of 19 days [52]. However, in contrast to our review, Morone et al. included more than 25 case reports of patients with frequently mild illness and/or follow-up periods that were not long enough to meet the inclusion critera of at least two weeks in our review. Both aspects may have led to a slightly shorter PR over time compared to our study.

In general, a common bias in larger studies is the focus on hospitalized patients, leading to an overrepresentation of more severe cases who stay in hospital for a longer time and might be more likely to shed virus longer in stool. Upon preparation of a review, this type of studies may potentially lead to a certain overestimation of shedding duration.

### Viral shedding dynamics: (b) quantity (viral load) over time (kinetics)

Due to the limited number of robust studies, only three studies were suitable to analyze the (quantitative) kinetics of the viral load [7,21,36]. Results suggest that viral shedding may peak during the first or early second week after symptom onset. This pattern applies to symptomatic individuals, as asymptomatic cases lack, by definition, a defined onset of symptoms. The observed time lag between peak shedding in the upper respiratory tract and stool—ranging from several days to about one week—may reflect the time needed for the virus to be swallowed, perhaps infect intestinal cells, and be excreted (Figure 5).

The three studies were conducted at different points in the pandemic, capturing periods with different dominant SARS-CoV-2 variants. We report normalized values, because the absolute quantities of viral load cannot be easily compared across variants due to differences in methodology and measurement. Notably, the duration of sample collection in one study spanned several consecutive periods with different dominant Omicron variants [21]. The authors noted that a significantly higher number of genome copies were measured for the BA.2 and JN.1 variants compared to those that preceded them chronologically or phylogenetically.

These findings have direct relevance for wastewater-based surveillance. The continued detectability of viral RNA in stool—despite vaccination, prior infections, and viral evolution—indicates that wastewater monitoring remains a feasible and informative tool. In addition, understanding shedding kinetics helps interpret wastewater signals more accurately—for example, in differentiating between ongoing transmission (incidence) and residual signal from prolonged shedding (prevalence). Clarifying this distinction is critical for accurate public health interpretation. Together, these insights support the continued value of wastewater surveillance in tracking SARS-CoV-2.

### Duration of SARS-COV-2 virus shedding in stool

Our review also investigated the maximum duration of SARS-CoV-2 viral shedding in stool across the included studies. Again, study methodologies varied widely. For many study investigators, systematic follow-up was a challenge—for example, when hospitalized patients were clinically fit for discharge but may still have had detectable viral RNA in their stool. Although many studies applied a systematic sampling approach—often defining the end of shedding after two consecutive negative stool samples—several others predefined a fixed period for sample collection regardless of patient shedding status. This design choice may have led to truncation of the actual shedding duration. For instance, Young et al. collected stool samples only during the first two weeks following study enrollment, with the maximum reported VST being 9 days [47]. Lo et al. followed participants for one month with a maximum VST of 19 days [32], while Vaselli et al. limited collection to three weeks, reporting a maximum VST of 21 days [44]. These predefined periods may have missed longer shedding durations. Nevertheless, it is noteworthy that two studies using a systematic sampling approach—defining the end of shedding after two consecutive negative stool samples— still reported relatively short maximum durations of shedding: 14 days [19] and 17 days [25]. Hence, because of the methodological differences, the range of reported maximum durations of fecal shedding (2-12 weeks) is not surprising. Interestingly, one study detected SARS-CoV-2 RNA in 4% of stool samples after 28 weeks [42]. Compared with the other studies this duration is exceptionally long. In this study, an average of 3 stool samples per patient suggests that patients have not closely been followed-up. Without longitudinal follow-up of the same patients, it is difficult to determine whether the detection of SARS-CoV-2 RNA in stool after such a long time reflects continuous shedding from a single infection or another, further infection acquired during the study period. Whole genome sequencing might have helped in validating the results. Regarding the question of the sequence of respiratory and fecal shedding, there seems to be a consensus that viral shedding in stool starts later and lasts longer than in the respiratory tract.

This review has a number of limitations. These include limited generalizability of our findings due to heterogeneity among the included studies and likely a substantial publication bias. We did not account for several factors that could be related to fecal viral shedding, such as demographic variables (e.g., sex and ethnicity), pre-existing conditions, and differences between symptomatic and asymptomatic individuals. We did take the vaccination status into consideration, however, as the vast majority of studies were conducted in 2020, vaccination was not an issue as vaccines were not available yet in 2020. Given the limited availability of high-quality studies, conclusive statements regarding differences in shedding kinetics among SARS-CoV-2 variants cannot be drawn.

For the research questions explored in this review, we propose a number of recommendations to enhance the quality of future studies, (Table 4).

**Table 4.**
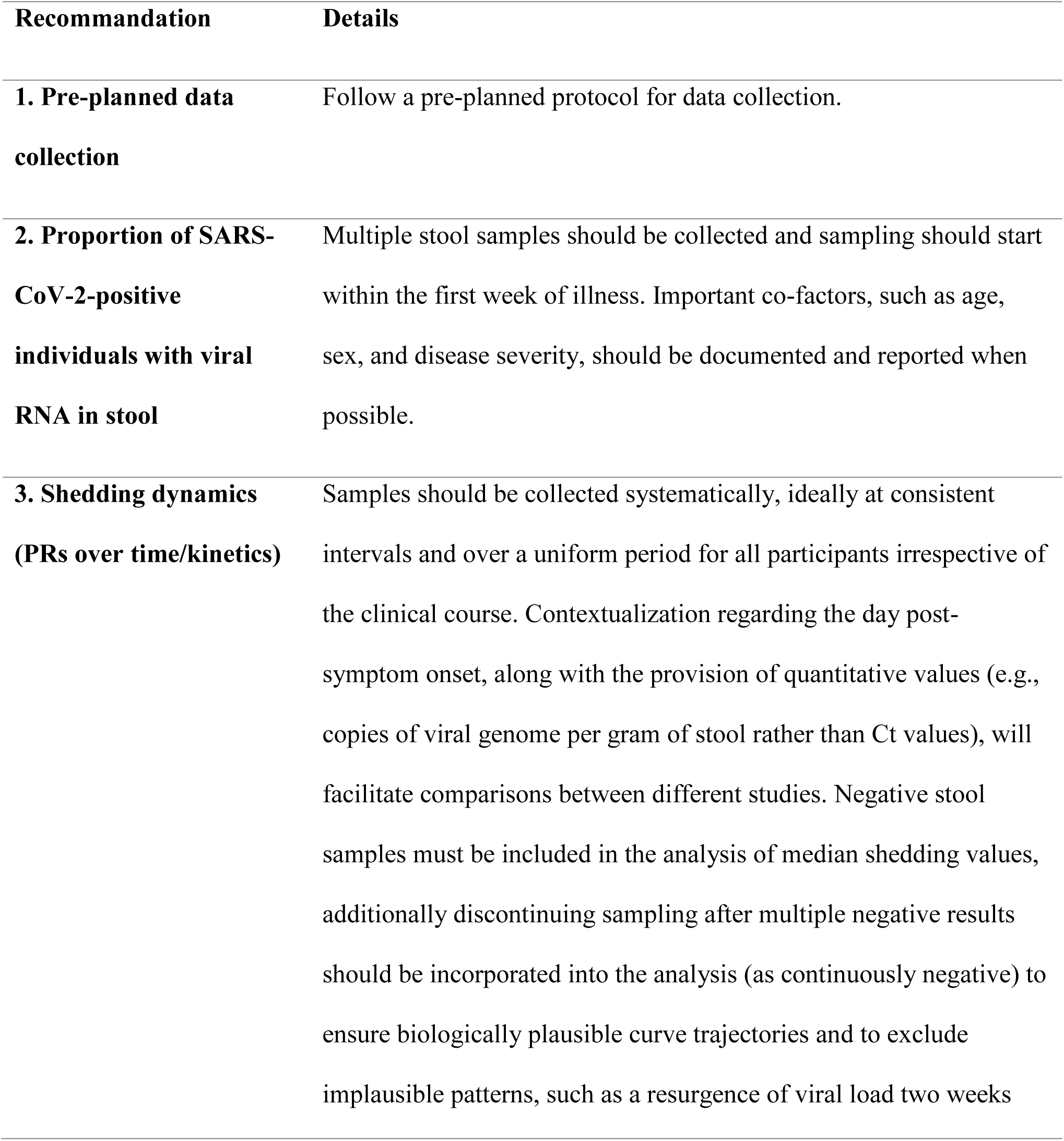
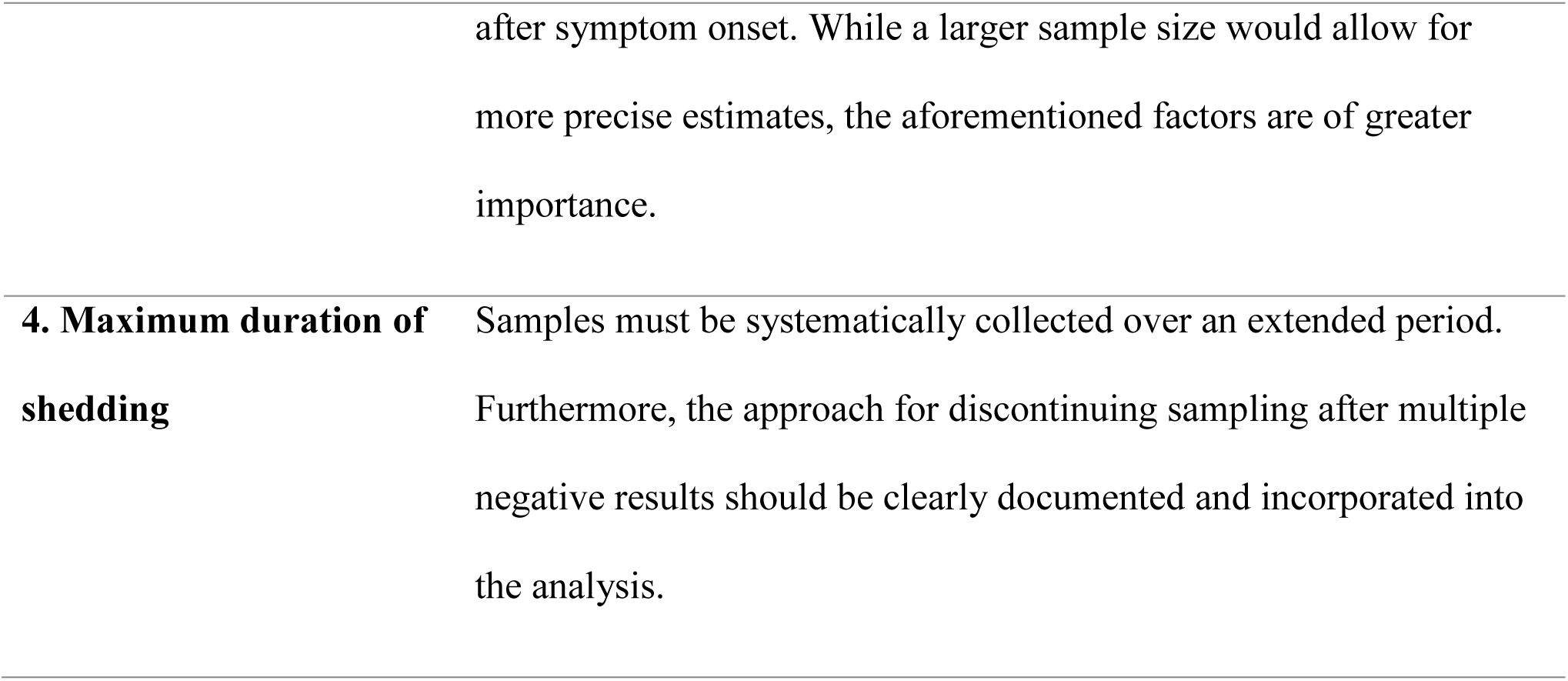
Recommendations for Enhancing the Quality of Future Studies on SARS-CoV-2 Stool Shedding.

## Conclusions

Our study suggests that viral shedding of SARS-CoV-2 can be detected in stool samples of approximately 50% of infected individuals. Among those with detectable viral shedding, the PR decreases to about 50% after three weeks post symptom onset, with viral load peaking between the third and ninth day after symptom onset. The maximum viral shedding duration may last up to 12 weeks. Methodological limitations, such as unsystematic data collection, unclear methodologies, and inadequate reporting of results, hinder the drawing of strong and reliable conclusions. The implementation of more standardized methods would enhance the robustness and comparability of findings.

## Data availability statement

The data from the scoping review is provided in the additional file. Further inquiries can be directed to the corresponding author.

## Abbreviations

PR: Positivity Rate
VST: Viral Shedding Time
CI: Confidence Interval
gc/g-wet: RNA gene copies per gram of wet stool
URT: Upper Respiratory Tract

## Ethics approval and consent to participate

Not applicable.

## Author contributions (CRediT taxonomy for contributors)

SA: Conceptualization, Data curation, Formal analysis, Investigation, Methodology, Project administration, Visualization, Writing – original draft, Writing – review & editing.

WH, JS: Conceptualization, Project administration, Writing – review & editing. TG, PP, AS: Methodology, Visualization, Writing – review & editing.

RK: Conceptualization, Data curation, Writing – review & editing.

UB: Conceptualization, Investigation, Methodology, Project administration, Formal analysis, Visualization, Writing – original draft, Writing – review & editing, Supervision.

