## Appendix for "Frequency, Dynamics, and Duration of Fecal Shedding in SARS-CoV-2 Infected Individuals, a Scoping Review"

**Table A1. Research Strategy for Fecal Viral Shedding of SARS-CoV-2-Infected Individuals.**

|  | PubMed |
| --- | --- |
| Population | COVID-19[MeSH] OR COVID-19[tiab] OR COVID19[tiab] OR "2019 novel coronavirus"[tiab] OR "2019 ncov"[tiab] OR "2019 ncov"[tiab] OR "coronavirus disease 2019"[tiab] OR "coronavirus disease-19"[ tiab] OR "severe acute respiratory syndrome coronavirus 2"[tiab] OR "SARS coronavirus 2"[tiab] OR SARS-CoV-2[tiab] |
| AND |  |
| Sample Material | stool[tiab] OR feces [MeSH] OR feces [tiab] OR faeces [tiab] OR fecal [tiab] |
| AND |  |
| Parameter | "Positivity rate"[tiab] OR duration[tiab] OR “detection rate”[tiab] OR "Virus Shedding"[MeSH] OR "shed*"[tiab] OR "viral kinetics"[tiab] OR "viral load kinetics"[ tiab] OR "viral dynamics"[tiab] OR "viral load dynamics"[tiab] OR "transmission kinetics"[tiab] OR "transmission dynamics"[tiab] OR "viral load"[tiab] OR "viral load"[MeSH] OR "Virus Shedding"[tiab] OR "temporal"[tiab] OR prevalence[tiab] OR prevalence[MeSH] |
| Limits | No limits were applied during this step |
| Output | 643 (18.July.2024) |

**Table A2: Summary of the metadata of the papers included in the review.**

| **Authors** | **Journal** | **Title** | **Study design** | **Participants´ number who provided stool samples** | **Countries involved/ setting** | **Time period of study execution** | **Participants**  **vaccinated*** |
| --- | --- | --- | --- | --- | --- | --- | --- |
| Vaselli et al. (2021) | BMC Infectious Diseases | Investigation of SARS-CoV-2 faecal shedding in the community: a prospective  household cohort study (COVID-LIV) in the  UK | Prospective cohort study | 17 | UK | 07.2020 –  09.2020 | No |
| Shi et al. (2020) | The Journal of Infectious Diseases | Clinical Characteristics and Factors Associated With  Long-Term Viral Excretion in Patients With Severe  Acute Respiratory Syndrome Coronavirus 2 Infection: a  Single-Center 28-Day Study | Retrospective cohort study | 99 | China | 01.2020 –  02.2020 | No |
| Lin et al. (2021) | Journal of Medial Virology | Association between detectable SARS‐COV‐2 RNA in anal swabs and disease  severity in patients with coronavirus disease 2019 | Prospective cohort study | 217 | China | 01.2020 –  02.2020 | No |
| Khemiri et al. (2023) | Frontiers in Medicine | SARS-CoV-2 excretion kinetics in nasopharyngeal and stool samples from the pediatric population | Prospective cohort study | 71 | Tunis | 02.2021 –  01.2022 | No |
| Deng et al. (2020) | BMC Infectious Diseases | Positive results for patients with COVID-19vdischarged form hospital in Chongqing,  China | Retrospective cohort study | 61 | China | 01.2020 –  03.2020 | No info. * |
| Pace et al. (2024) | Frontiers in Immunology | Prevalence and duration of SARS-CoV-2 fecal shedding in breastfeeding dyads following maternal COVID-19 diagnosis | Retrospective cohort study | 61 | USA | 04.2020 –12.2021 | One vaccinated participant out of the 61 participants |
| Wang et al. (2020) | Virus Research | Fecal viral shedding in COVID-19 patients: Clinical  significance, viral load  dynamics and survival analysis | Retrospective cohort study | 69 | China | 02.2020 –  01.2020 | No info. |
| De Ioris et al. (2020) | Journal of the Pediatric Infectious Diseases Society | Dynamic Viral Severe Acute  Respiratory Syndrome  Coronavirus 2 RNA  Shedding in Children:  Preliminary Data and  Clinical Consideration from  a Italian Regional Center | Prospective cohort study | 22 | Italy | 03.2020 –  04.2020 | No info. |
| Zhang S et al. (2021) | Journal of Clinical Laboratory Analysis | Risk factors for prolonged virus shedding of respiratory tract and fecal in adults with severe acute respiratory syndrome coronavirus-2 infection | Retrospective cohort study | 126 | China | 01.2020 –  04.2020 | No info. |
| Natarajan et al. (2022) | Med | Gastrointestinal symptoms and fecal shedding of SARS-CoV-2 RNA suggest prolonged  gastrointestinal infection | Prospective cohort study | 102 | USA | 04.2020 – 07.2020 | No info. |
| Young et al. (2020) | JAMA | Epidemiologic Features and Clinical Course of Patients Infected With SARS-CoV-2 in  Singapore | Prospective cohort study | 8 | Singapore | 01-2020 –  02.2020 | No info. |
| Holm-Jacobsen et al. (2021) | Frontiers in Medicine | The Prevalence and Clinical Implications of Rectal SARS-CoV-2 Shedding in Danish COVID-19 Patients and the General Population | Prospective cohort study | 52 | Denmark | 06.2020 – 02.2021 | One vaccinated participant out of the 52 participants |
| Wu Y et al. (2020) | The Lancet | Prolonged presence of SARS-CoV-2 viral RNA in faecal samples | Prospective cohort study | 74 | China | 01.2020 –  03.2020 | No info. |
| Zheng et al. (2020) | The BMJ | Viral load dynamics and disease severity in patients infected  with SARS-CoV-2 in Zhejiang province, China, January-March  2020: retrospective cohort study | Retrospective cohort study | 93 | China | 01.2020 –  03.2020 | No info. |
| Lavania et al. (2022) | Frontiers in Medicine | Prolonged Shedding of SARS-CoV-2 in Feces of COVID-19 Positive Patients: Trends in Genomic Variation in First and Second Wave | Prospective cohort study | 280 | India | 05.2020 –  08.2021 | No info. |
| Chen et al. (2020) | Journal of Medial Virology | The presence of SARS‐CoV‐2 RNA in the feces of COVID‐19  patients | Prospective cohort study | 42 | China | 01.2020 –  02.2020 | No info. |
| Kujawski et al. (2020) | Nature Medicine | Clinical and virologic characteristics of the first  12 patients with coronavirus disease 2019  (COVID-19) in the United States | Case Series | 10 | USA | 01.2020 –  02.2020 | No info. |
| Jung et al. (2021) | Gastroenterology &  Hepatology | Serial Screening for SARS-CoV-2 in Rectal Swabs of Symptomatic COVID-19  Patients | Prospective cohort study | 10 | South Korea | No info. But the study was received and accepted in 2021 | No info. |
| Mesoraca et al. (2020) | Virology Journal | Evaluation of SARS-CoV-2 viral RNA in fecal samples | Case series | 15 | Italy | 03.2020 –04.2020 | No info. |
| Arts et al. (2023) | mSphere | Longitudinal and quantitative fecal shedding dynamics of SARS-CoV-2, pepper mild mottle virus, and crAssphage | Prospective cohort study | 48 | USA | 09.2020 –  04.2021 | Three of the four vaccinated individuals included in this cohort had measurable SARS-CoV-2 RNA in their stool at some point in the sampling period |
| Yuan et al. (2021) | PLoS ONE | SARS-CoV-2 viral shedding characteristics and potential evidence for the priority for faecal specimen testing in diagnosis | Prospective cohort study | 10 | China | 01.2020 –  03.2020 | No info. |
| Zhang N et al. (2021) | Science China Life Sciences | Comparative study on virus shedding patterns in nasopharyngeal and fecal specimens of COVID-19 patients | Retrospective cohort study | 12 | China | 01.2020 –  02.2020 | No info. |
| Wu B et al. (2020) | Infectious Diseases of Poverty | Compare the epidemiological and clinical features of imported and local COVID-19  cases in Hainan, China | Retrospective cohort study | 91 | China | 01.2020 –  02.2020 | No info. |
| Zuo et al. (2020) | Gastroenterology | Alterations in Gut Microbiota of Patients With COVID-19 During Time of Hospitalization | Prospective cohort study | 15 | China | 02.2020 –  03.2020 | No info. |
| Liu et al. (2020) | Emerging Microbes & Infections | Dynamic surveillance of SARS-CoV-2 shedding and neutralizing antibody in  children with COVID-19 | Retrospective cohort study | 9 | China | 01.2020-02.2020 | No info. |
| Wölfel et al. (2020) | Nature | Virological assessment of hospitalized patients with COVID-2019 | Prospective cohort study | 9 | Germany | 01.2020 | No info. |
| Hua et al. (2020) | Journal of Medial Virology | Epidemiological features and viral shedding in children with  SARS‐CoV‐2 infection | Retrospective cohort study | 35 | China | 02.2020 | No info. |
| Cai et al. (2020) | Virologica Sinica | Comparison of Clinical and Epidemiological Characteristics  of Asymptomatic and Symptomatic SARS-CoV-2 Infection in Children | Prospective cohort study | 49 | China | 01.2020 – 04.2020 | No info. |
| Han et al. (2020) | Emerging Infectious Diseases | Viral RNA Load in Mildly  Symptomatic and  Asymptomatic Children  with COVID-19, Seoul,  South Korea | Prospective cohort study | 12 | South Korea | 03.2020 –04.2020 | No info. |
| Jeong et al. (2020) | Clinical Microbiology and Infection | Viable SARS-CoV-2 in various specimens from COVID-19 patients | Prospective cohort study | 5 | South Korea | 02.2020 –  03.2020 | No info. |
| Lo et al. (2020) | International Journal of Biological Sciences | Evaluation of SARS-CoV-2 RNA shedding in clinical  specimens and clinical characteristics of 10 patients with  COVID-19 in Macau | Prospective cohort study | 10 | China | 01.2020 | No info. |
| Wannigama et al. (2024) | The Lancet | Increased faecal  shedding in SARS-CoV-2 variants BA.2.86 and  JN.1 | Prospective cohort study | 113 | Japan | 09.2023 –  12.2023 | Yes;  “The cohort contained  all individuals (n=113) fully  vaccinated for SARS-CoV-2, with  eight (7%) having received BA4/5  bivalent booster.“ |
| Sun et al. (2020) | Emerging Infectious Diseases | Prolonged Persistence of  SARS-CoV-2 RNA in Body Fluids | Prospective cohort study | 49 | Chin | In 2020 | No info. |

*No info.; no data available about the vaccinated participants, as most studies were conducted in 2020 and early 2021 when vaccination was not widely available

**Table A3: Maximum viral shedding duration of SARS-CoV-2 RNA in stool across studies that adopted a systematic sampling approach over the follow-up period. N: the total number of participants with detectable virus in stool samples included in the study. n1: the number of participants who tested positive on the day of maximum viral shedding time (VST). n2: The total number of participants tested** **on the day of the maximum VST. Max VST: The maximum duration of viral shedding time in days. NA: Not available.**

| Author | N | n1/n2 | n2/N | Max VST | Note |
| --- | --- | --- | --- | --- | --- |
| Sun et al. (2020) | 49 | NA | NA | 49 | Provided the estimated 95% percentile using a Weibul model. Feces specimens were collected every 3 days for 4 weeks until having a negative result. |
| Lavania et al. (2022) | 173 | NA | NA | 55 | n2 is not defined. Fecal samples from COVID-19 patients were collected twice during hospitalization, at discharge, and monthly if still positive. |
| Zheng et al. (2022) | 55 | 1/NA | NA | 60 | n2 is not defined. Stool samples were collected daily whenever possible. |
| Arts et al. (2023) | 35 | 2/11 | 11/35 | 28 | Stool samples were collected daily whenever possible. |
| Cai et al. (2020) | 45 | NA | NA | 70 | n2 is not defined. Samples were collected every three days until SARS-CoV-2 was undetectable. |
| De Ioris et al. (2020) | 7 | NA | NA | 17 | n2 is not defined. Samples were repeated every 2–3 days until there were 2 consecutive negative results. |
| Holm-Jacobsen et al. (2021) | 12 | 1/1 | 1/12 | 45 | Patients were tested until having two consecutive negative tests. The patient who provided the last positive test had two negative sample after the last positive test. Another patient was tested after this time point and was also negative. |
| Mesoraca et al. (2020) | 11 | 4/11 | 11/11 | 30-40 | The fecal specimens were collected every 5 days until the final date of the study. |
| Pace et al. (2024) | 17 | 3/NA | NA | During week 4 | n2 is not defined. Most of the patients provided samples during the eighth week, and all of them tested negative. Fecal samples were collected on days 1, 2–6, 7, and at 2, 3, 4, and 8 weeks post-enrollment. |
| Wang X et al. (2020) | 20 | 1/NA | NA | 49 | n2 is not defined. SARS-CoV-2 RNA was tested in stool until two sequential negative results were obtained. |
| Wu Y et al. (2020) | 41 | 1/NA | NA | 50 | n2 is not defined. Fecal samples were collected every 1–2 days until two sequential negative results were obtained. |
| Zhang S et al. (2021) | 60 | 1/2 | 2/60 | 85 | Viral shedding duration was measured from symptom onset until two consecutive negative tests. Two patients were tested on day 90 after symptom onset, one tested positive. Both patients achieved the condition of having two consecutive negative samples after day 90. |
| Khemiri et al. (2023) | 15 | 6/15 | 15/15 | 14 | Samples were collected on a regular basis from the onset of the infection until having a negative result. Six patients were tested after the day of the maximum VST and were negative. |

**Table A4: Maximum viral shedding duration of SARS-CoV-2 RNA in stool across studies that did not provide information on or did not adopt a systematic sampling approach over the follow-up period. N: the total number of participants with detectable virus in stool samples included in the study. n1: the number of participants who tested positive on the day of maximum viral shedding time (VST). n2: The total number of participants tested on the day of the maximum VST. Max VST: The maximum duration of viral shedding time in days. NA: Not available.**

| Author | N | n1/n2 | n2/N | Max VST | Note |
| --- | --- | --- | --- | --- | --- |
| Wölfel et al. (2020) | 9 | 1/2 | 2/26 | 26 |  |
| Lo et al. (2020) | 10 | 1/3 | 3/10 | 19 |  |
| Han et al. (2020) | 11 | 1/1 | 1/11 | 27 |  |
| Kujawski et al. (2020) | 7 | 1/1 | 1/7 | 25 | Two samples were taken from another two patients after 25 day of symptom onset  and were negative. |
| Zhang N et al. (2021) | 10 | 1/3 | 3/10 | 26 |  |
| Hua et al. (2020) | 32 | 1/NA | NA | >70 | n2 is not defined. |
| Lin et al. (2021) | 46 | 2/9 | 9/46 | 38-40 | Seven patients provided samples after days 38-40 after symptom onset and all were negative. |
| Liu et al. (2020) | 8 | 1/1 | 1/8 | 66 | The last patient with a positive stool on day 66 after symptom onset provided another sample on day 70 and tested negative. |
| Vaselli et al. (2021) | 3 | 1/2 | 2/3 | 21 |  |
| Natarajan et al. (2022) | 50 | 2/NA | NA | 210 | Exact n2 could not be determined from the study data; 60 patients provided samples, including those with and without detectable virus. A total of 673 stool samples were collected from 113 participants over the study period. |
| Young et al. (2020) | 4 | 1/2 | 2/4 | 9 | On day 13, stool samples were provided by only three patients, all of which tested negative. |
| Zuo et al. (2020) | 13 | 1/NA | NA | 33 | n2 is not defined. |
